# Improved access to diabetic retinopathy screening through primary care-based teleophthalmology during the COVID-19 pandemic

**DOI:** 10.1101/2023.05.03.23289435

**Authors:** Eliot R. Dow, Karen M. Chen, Marina Basina, Jimmy Dang, Nergis C. Khan, Michael Kim, Marcie Levine, Kapil Mishra, Chandrashan Perera, Anuradha Phadke, Marilyn Tan, Kirsti Weng, Diana V. Do, Vinit B. Mahajan, Prithvi Mruthyunjaya, Theodore Leng, David Myung

## Abstract

**Background:** Primary care practices play a critical role in ensuring that patients with diabetes undergo an annual eye examination, the importance of which is underscored by the Healthcare Effectiveness Data and Information Set (HEDIS) quality measures. Store-and-forward teleophthalmology, where ocular images are read remotely by an ophthalmologist, has the potential to facilitate this role.

**Methods:** In this report, we aim to measure if using a primary care-based teleophthalmology program improves access to eye examinations for diabetic patients as reflected in HEDIS measures. Over a 20-month period, non-mydriatic fundus photographs were obtained at five primary care sites in the San Francisco Bay Area from patients with a new or existing diagnosis of diabetes mellitus type 1 or 2 who needed an annual eye examination. Collected photographs were evaluated remotely by vitreoretinal specialists for diabetic retinopathy. Our primary measures were the proportion and number of annual eye exams of diabetic patients in primary care clinics that participated in the teleophthalmology program compared to clinics that did not participate. Additional measures included the number of patients with DR who were identified through the program, gradeability of fundus photographs, and characteristics of the study population.

**Results:** The program screened 760 unique patients, 84 of whom were found to have DR (11.1%). The rate of ungradable photos was 9.7%, which was greater for patients who self-reported as racially non-White. For the duration of the study, including during the COVID-19 pandemic, both the proportion and number of diabetic patients receiving annual eye examination increased (17.1% increase in proportion, 14.8% increase in number). In comparison, primary care sites that did not offer the teleophthalmology service declined in these measures (2.3% decrease in proportion, 17.0% decrease in number).

**Conclusions:** Primary care-based teleophthalmology improves access to eye exam for diabetic patients and identifies patients with diabetic retinopathy across diverse communities.

## BACKGROUND

Eye diseases worldwide, including within the United States, are underdiagnosed and undertreated^1^. A multitude of factors contribute to this deficiency in eye care including, but not limited to, availability of specialists, transportation and mobility barriers, financial burden, lack of education, and poor patient-physician communication and understanding^2,3,4^.

Teleophthalmology, a paradigm of care delivery in which ocular images are interpreted remotely by an eye specialist, has increased in interest since the COVID-19 pandemic, may offer improved access to necessary eye care^5^.

The need for improved access through teleophthalmology is particularly critical for diabetic retinopathy (DR), the leading cause of new cases of blindness among adults aged 20 to 60 affecting more than 100 million patients worldwide^6,7^. DR arises when elevated levels of blood sugar resulting from either type 1 or 2 diabetes mellitus damage the blood vessels that supply oxygen and nutrients to the retina, the light-sensing part of the eye. The risk of developing DR is directly related to the length of time that a patient has diabetes and usually does not appear for approximately five years after a type 1 diabetes diagnosis, although it may already be present when type 2 diabetes is diagnosed^8^. In the absence of glycemic control and/or ophthalmic treatment, the disease may progress through three stages of non-proliferative retinopathy (mild, moderate, severe) before proliferative retinopathy develops. Diabetic macular edema can occur at any stage of retinopathy. If DR is diagnosed early, vision loss may be mitigated or prevented^9^. An annual fundus examination to screen for DR is critical, however, only about half of all patients with diabetes receive proper screening and less than 40% of patients with a high risk of vision loss ever undergo treatment^10,11^.

In 2010, primary care providers (PCPs) delivered clinical care to approximately 90% of individuals with type 2 diabetes, and the proportion has increased over time^12^. The importance of primary care practitioners ensuring that their diabetic patients receive recommended eye care is reflected in the Healthcare Effectiveness Data and Information Set (HEDIS). This comprehensive set of quality performance measures across six domains of care guide the primary care of chronic medical conditions like diabetes mellitus and includes assessment of whether a diabetic patient receives diabetic eye screening at least every two years^13^. Attainment of these quality measures is increasingly important for health-system quality ratings and value-based reimbursement models.

Practices are increasingly turning to teleophthalmology programs to aid in this goal of care^5,14^. Traditionally, DR is diagnosed by an eye specialist via an annual in-person fundoscopic examination. However, with appropriate training, non-ophthalmic clinicians and clinical personnel are able to use a fundus camera to take retinal photos that can then be evaluated by an ophthalmologist typically via a store-and-forward model. DR can be determined with high sensitivity and specificity from fundus photography, and a referral for further ophthalmic evaluation or treatment is made for those patients with retinopathy^15^. Primary care-based teleophthalmology programs have improved the accessibility and cost-effectiveness of DR screening in both rural and urban settings worldwide and are currently being applied to DR screening more commonly than any other ocular pathology^16, 17,18^. The ongoing COVID-19 pandemic has exacerbated existing barriers and increased the likelihood of ophthalmic appointment postponement or cancellation rendering teleophthalmology services even more critical to DR screening programs^19, 20^.

The prevalence of diabetes in California is more than 40% above the United States national average^21^. As a means to improve the ophthalmic health of our patients, the Stanford Teleophthalmology Automated Testing and Universal Screening (STATUS) program was developed as a multi-site teleophthalmology DR screening collaboration between the Byers Eye Institute of Stanford (BEIS) and five affiliated primary care clinics throughout the San Francisco Bay Area. The program was initiated two to six months (depending on the site) prior to the onset of the COVID-19 pandemic in the United States and continued to provide remote eye examinations to patients throughout 2020 and 2021. The goal of the program was to evaluate whether the use of teleophthalmology could increase the percentage of patients screened for DR in collaboration with regional primary care clinics. Here, we examine the ability of the 18-month teleophthalmology program to improve and maintain access to DR eye care prior to and during the COVID-19 pandemic.

## METHODS

### Clinic Sites

Non-mydriatic fundus cameras were deployed at an academic-affiliated primary care site in Santa Clara, CA in September 2019, and in four additional affiliated primary care sites in Los Gatos, Oakland, Hayward, and Pleasanton, CA beginning in February 2020. The primary care sites ranged from 20 miles (25-minute drive) to 42 miles (45-minute drive) away from the BEIS. Store-and-forward teleophthalmology screening for diabetic retinopathy continued at all five locations throughout the study period which ended April 2021. In order to determine whether the teleophthalmology program impacted the adherence rate to annual diabetic eye exams, HEDIS measures at two primary care sites (Pinole, CA and San Pablo, CA) in the same healthcare system that did not deploy the teleophthalmology system were also assessed. The study was approved by the Institutional Review Board at Stanford University.

### Patient Image Collection and Assessment

Patients 18 years or older with type 1 or type 2 diabetes mellitus without a prior DR diagnosis or a DR exam in the past 12 months were offered the opportunity to have fundus photographs taken at the end of their primary care visit. Fundus imaging was performed by a trained medical assistant using the CenterVue DRS fundus camera (Hillrom Inc., Chicago, IL) at the Santa Clara primary clinic site and the TopCon NW400 fundus camera (Welch Allyn Inc., Skaneateles Falls, NY) at the Los Gatos, Oakland, Hayward, and Pleasanton primary care clinics. If medical assistants deemed the image quality to be poor, they repeated image acquisition and did so up to 4 times. The fundus images were forwarded to vitreoretinal specialists at BEIS who evaluated the images within one week. These fundus images were classified as ungradable (such as when opacity, blurring, or decentration impaired visualization of the fundus), or gradable if the quality was sufficient for grading of DR. Images of adequate quality had a DR grade assigned in accordance with the International Clinical Diabetic Retinopathy Disease Severity Scale with moderate and severe categories combined on teleophthalmology evaluation (no diabetic retinopathy/mild non-proliferative diabetic retinopathy/moderate to severe non-proliferative diabetic retinopathy/proliferative diabetic retinopathy)^22^. Patient images were also assessed for the presence of macular edema or other fundus abnormalities. Patients with images of insufficient quality from one or both eyes were recommended to have the images retaken or present for an in-person eye examination. Diagnosis and stage of DR were determined by the eye with more advanced retinopathy. Those with referral-warranted disease were referred for an in-person exam at BEIS or their local ophthalmologist. A subset of patients (N=26) voluntarily presented for a second teleophthalmology screening one year after their first examination.

### Patient Data

Patient files containing information on labs, orders, clinical notes, and patient information were retrieved from The STAnford Research Repository (STARR), an institutional resource for working with clinical data for research purposes. Data was managed and analyzed using Python (version 3.9.0) with Pandas (version 1.3.0). Patients who underwent fundus imaging without a documented assessment by BEIS specialists were excluded (N = 23). For all patients who were seen at BEIS after a referral for in-person examination, data was manually collected from the electronic health record. For analyses comparing patients prior to and during the COVID-19 pandemic, March 16th, 2020, was used as the start of the pandemic since on that date legal stay-at-home orders were announced in Alameda, Contra Costa, Marin, San Francisco, San Mateo, and Santa Clara counties. Longitudinal HEDIS data were only available for three of the teleophthalmology primary care sites and the two non-teleophthalmology comparison sites; two teleophthalmology primary care sites did not have structured HEDIS data available for analysis.

## RESULTS

Over the period from September 2019 to April 2021, 760 unique patients with diabetes mellitus participated in the teleophthalmology system, for a total of 790 screenings done. The median age of participating patients was 60.0 years (range 25-99), and 45.3% were women. The study population was 32.0% White, 25.6% Asian, 8.6% Black, 2.3% Native American / Pacific Islander, and 31.5% Other / Unknown based on self-reported race. Additionally, the study population was 21.9% Hispanic, 69.2% Non-Hispanic, and 8.9% Other / Unknown ethnicity by self-report (Figure 1). The teleophthalmology system provided service to racially and ethnically diverse communities within the five primary care sites, each having a different majority self-reported race or ethnicity (Supplemental Figure 1.).

**Figure 1.**
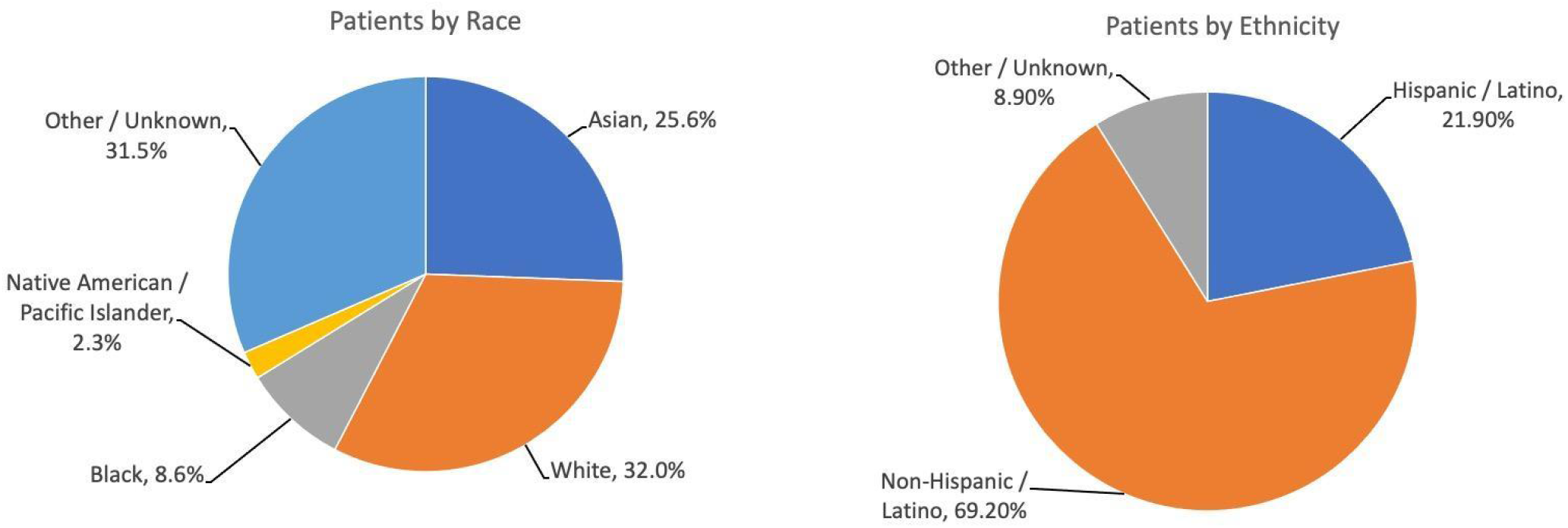
Self-reported race (top) and ethnicity (bottom) of patients participating in teleophthalmology program

Images were of sufficient quality to assess DR status in 713 of 790 screens (90.3%). The ungradable images (N=77) were significantly more likely to be obtained from older patients (68.1 years for ungradable vs. 58.5 years for gradable; p < 0.0001. Additionally, patients who self-reported as ethnically non-White were also more likely to have ungradable images (White 5.5%, Non-White 13.3%. Chi-squared 7.5107; p < 0.006) (Figure 2). This trend could not be accounted for by differences in age since non-White patients were, on average, a decade younger than White patients (76.9 years for White vs. 66.1 for non-White; p < 0.002).

**Figure 2.**
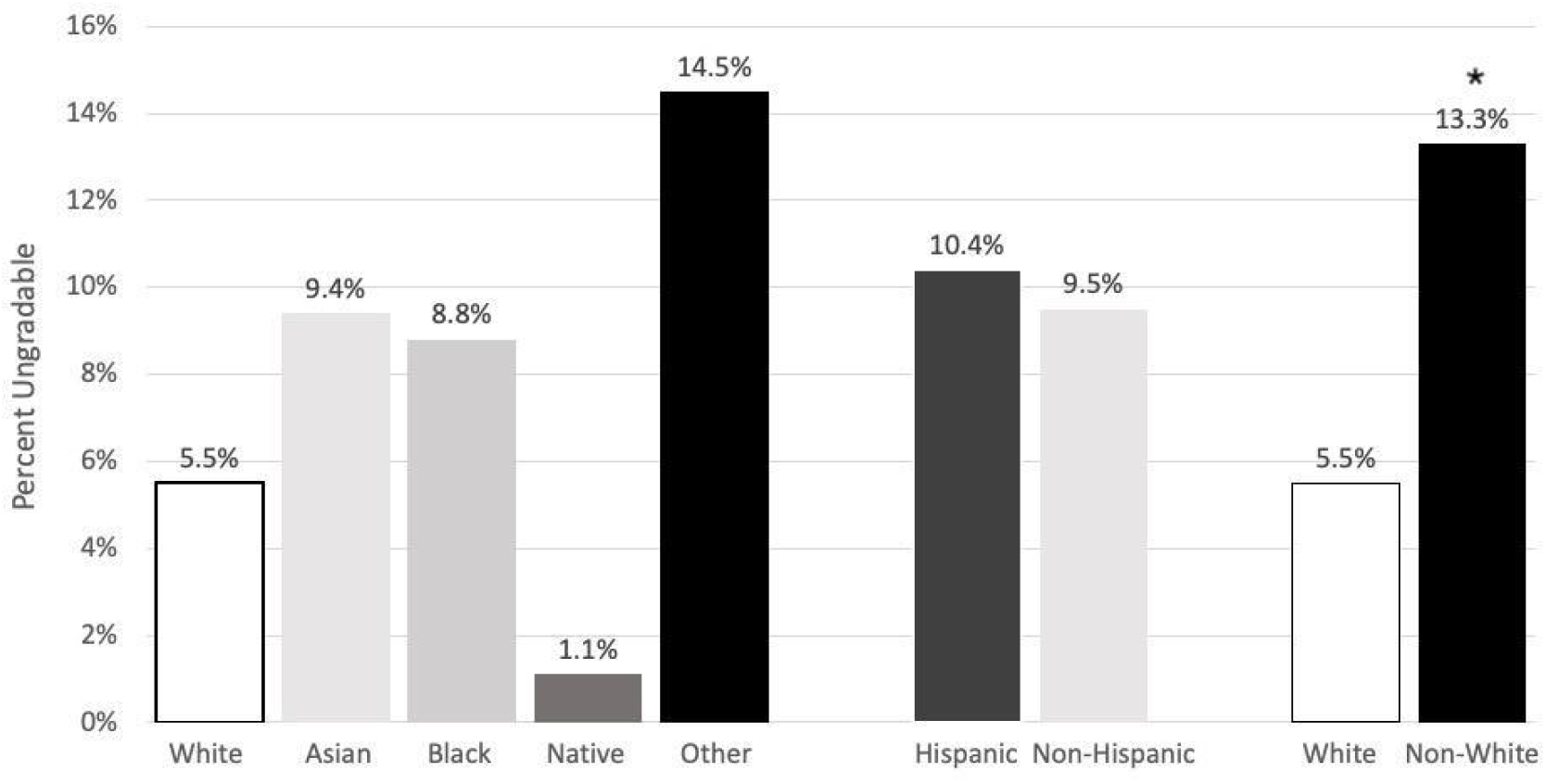
Self-reported race or ethnicity of patients with ungradable fundus photographs

The screening program detected 84 patients with diabetic retinopathy (11.1%), of whom 6.2% had mild DR, 4.7% had moderate or severe non-proliferative DR, and 0.1% have proliferative DR. In our study, patients diagnosed with any DR tended to be younger than those without DR (median age 54.1 years vs. 59.1 years, p < 0.001). Patients with and without DR did not differ in the likelihood of having hypertension or cardiovascular disease or in their glucose, Hemoglobin A1c, LDL, or HDL levels (Table 1). Among 26 patients who voluntarily presented for consecutive annual teleophthalmology examinations, none progressed in their stage of retinopathy.

**Table 1.** Systemic medical comorbidities and laboratory values of patients screening positive for diabetic retinopathy compared to those screening negative for diabetic retinopathy.

| Characteristic | DR (positive) | No DR (negative) | P-value |
| --- | --- | --- | --- |
| Age (years) | 54.1 | 59.1 | < 0.001 |
| Hypertension (% with comorbidity) | 89.4% | 87.2% | 0.86 |
| Cardiovascular Disease (% with comorbidity) | 84.7% | 83.9% | 0.94 |
| Average Last Glucose Level (mg/dL) | 170.1 | 158.2 | 0.106 |
| Average Last Hemoglobin A1c Level (%) | 9.02% | 8.83% | 0.298 |
| Average Last LDL Level (mg/dL) | 95.5 | 94.4 | 0.81 |
| Average Last HDL Level (mg/dL) | 53.4 | 52.4 | 0.431 |

The 84 patients positive for diabetic retinopathy on screening as well as the 77 patients with ungradable images were referred for an in-person eye examination. All patients were allowed to follow up at an eye care provider of their choice. Of these 161 patients, 18 patients presented for follow-up at BEIS (11.2% of those referred). Data was not available regarding the completion or location of follow-up for the remainder of the referred patients who were seen outside of the Stanford system. The patients who followed up at BEIS were seen a median of 14.5 days (S.D. ∓ 29.9 days) after referral for exam. On in-person examination, patients were found to have the same DR stage as the diagnosis made by teleophthalmology in 9 out of 17 eyes (46.7%), however, this increased to 14 of 17 eyes (82.4%) when analyzed as the presence versus absence of DR (Figure 3). In terms of diabetic macular edema, the teleophthalmology diagnosis was concordant with the in-person diagnosis in 9 out of 17 eyes (56.7%), which included both false positive and false negative cases. For patients with ungradable images who followed up at BEIS, 12 out of 14 eyes had no DR or DME (Figure 3).

**Figure 3.**
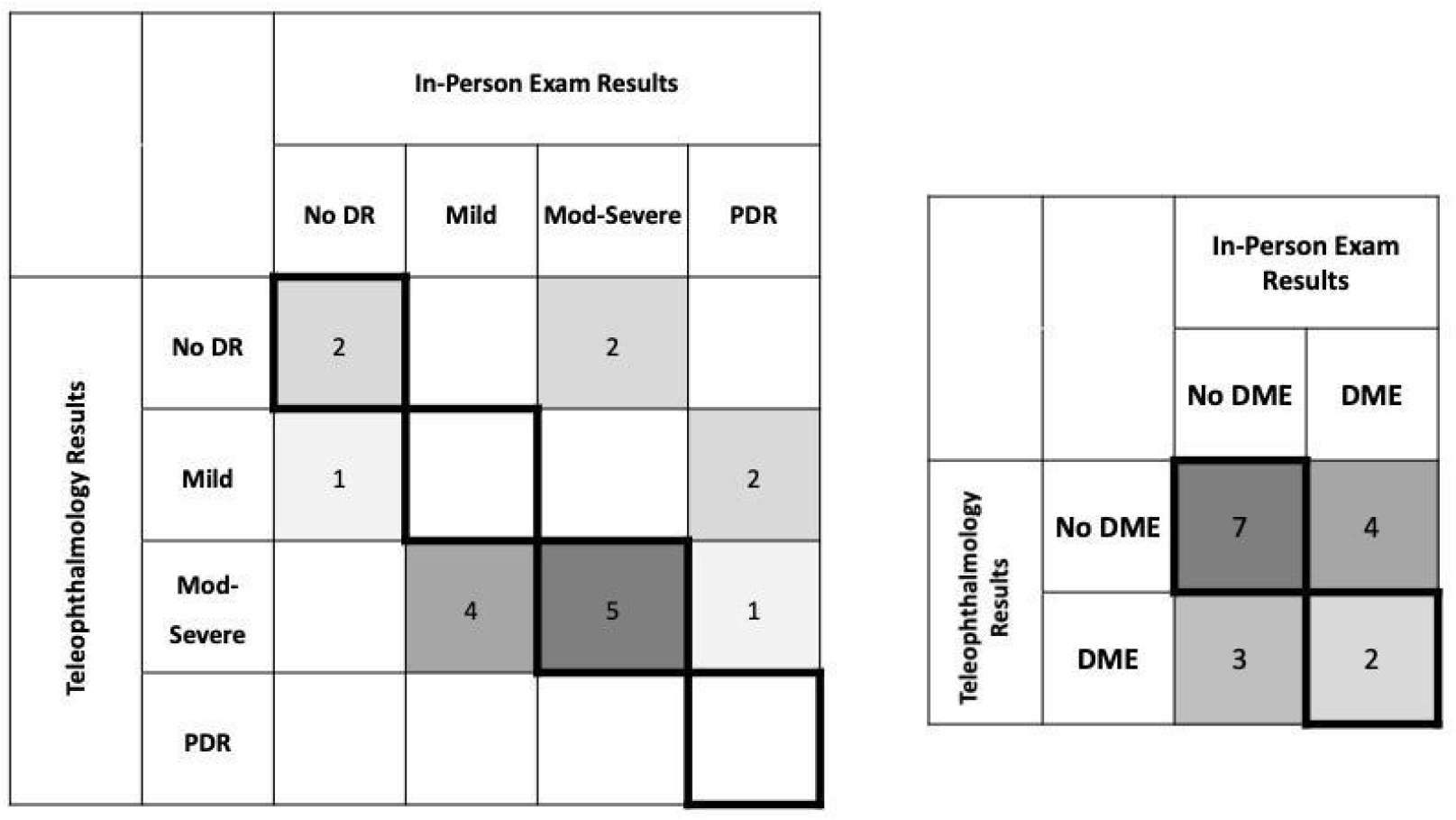
Confusion matrices comparing diagnosis of diabetic retinopathy and diabetic macular edema by teleophthalmology (y-axis) versus in-person examination (x-axis).

In accordance with the HEDIS measure specification for Comprehensive Diabetes Care, the primary care clinics tracked the percentage of the diabetic patient population up-to-date with recommended diabetic eye exams. A target of 67.89% was chosen, reflecting the 90th percentile HEDIS national benchmark. The percentage of patients who were adherent with annual diabetic eye exams increased from an average baseline of 49.5% across the study sites prior to implementation of teleophthalmology to an average of 66.6% across all months of the teleophthalmology system with 73.2% of patients receiving the recommended diabetic eye exam in the final month of the study period. For comparison, two primary care sites within the same health care system that did not implement the teleophthalmology system slightly decreased during the same interval of time from 72.6% of eligible patients receiving eye exams to 70.3% receiving them (Figure 4A). In terms of the absolute number of diabetic patients who received their eye exam, the primary care sites that implemented teleophthalmology had on average 14.8% more patients completing a diabetic eye exam after teleophthalmology was implemented compared to before. In contrast, the primary care sites without teleophthalmology decreased in the number of patients screened for diabetic eye disease by 17.0% during the same interval (Figure 4B). Although we had hypothesized that patients with systemic cardiovascular comorbidity and poorly controlled diabetes would be less likely during the COVID-19 pandemic to present for diabetic eye examination due to an exacerbation of care barriers, there was no significant difference between those populations for a variety of diseases or disease severity markers (Table 2).

**Table 2.** Systemic medical comorbidities and laboratory values of diabetic patients participating in teleophthalmology system before and after the COVID-19 pandemic.

| Characteristic | Pre-COVID | Post-COVID | P-value |
| --- | --- | --- | --- |
| Age (years) | 60.2 | 60.6 | 0.72 |
| Hypertension (% with comorbidity) | 89.75% | 86.9% | 0.83 |
| Cardiovascular Disease (% with comorbidity) | 89.4% | 84.4% | 0.61 |
| Average Last Glucose Level (mg/dL) | 166.3 | 157.6 | 0.100 |
| Average Last Hemoglobin A1c Level (%) | 8.62% | 8.92% | 0.126 |
| Average Last LDL Level (mg/dL) | 94.5 | 97.9 | 0.83 |
| Average Last HDL Level (mg/dL) | 50.9 | 53.0 | 0.302 |

**Figure 4.**
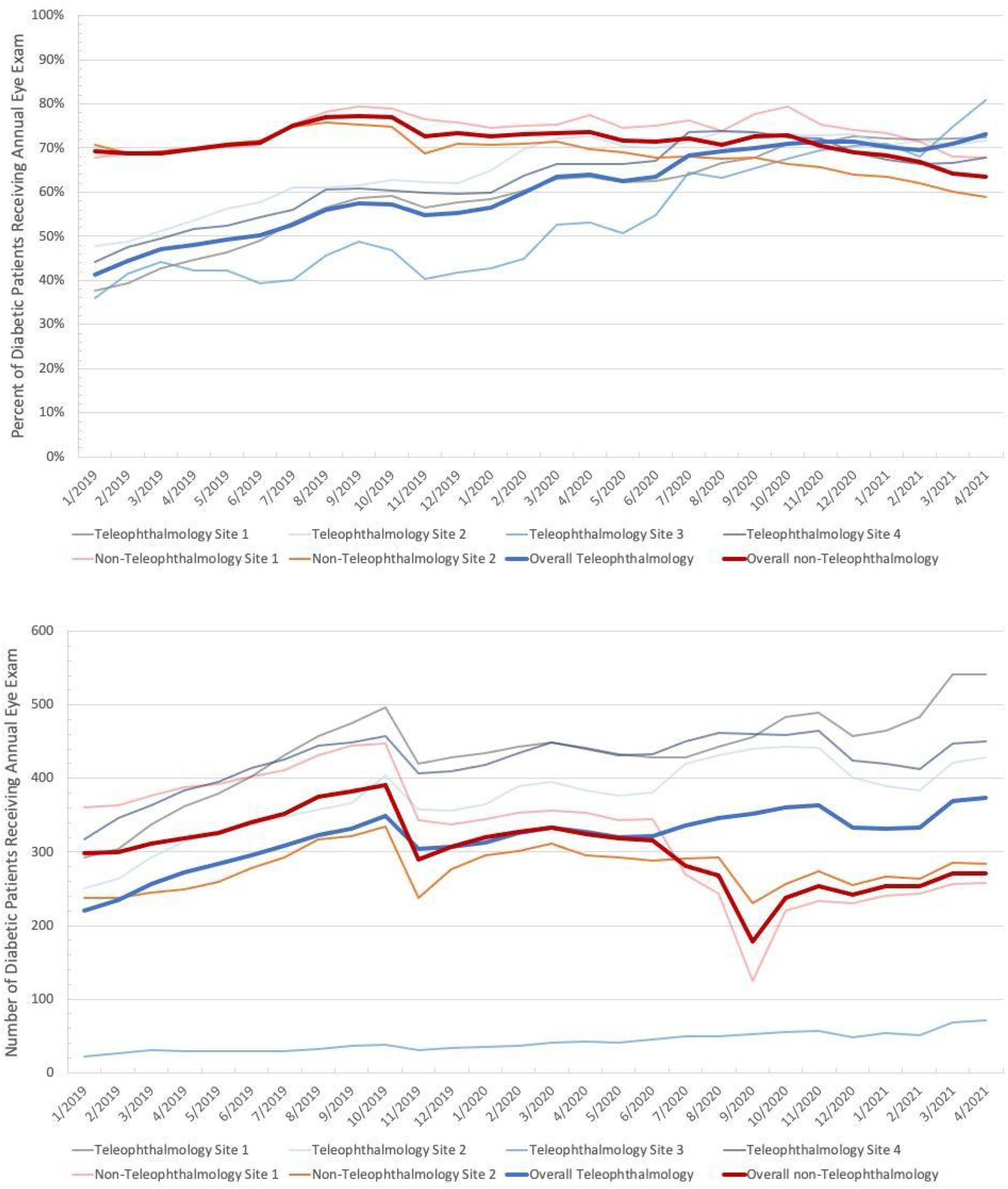
Percentage (top) or number (bottom) of patients receiving diabetic eye examination at sites offering teleophthalmology (blue lines) versus sites not offering teleophthalmology (red lines).

## DISCUSSION

Here we report on a teleophthalmology system for DR screening using fundus photography implemented at five primary care sites in the San Francisco Bay Area across 20 months. The program detected DR in 11.1% of screened patients, a figure that is lower than previously reported rates^23^. This may reflect true differences in prevalence among the patient population of the Bay Area compared to other populations. The laboratory values seen in our patient population represent relatively controlled diabetes, and thus, the system we used may be even more valuable in patients from rural and low socioeconomic areas with poor blood sugar maintenance. This may be particularly attractive for low-resource settings, since the program was carried out by trained medical assistants without the need for hiring additional staff.

Alternatively, the discrepancy could represent a lower sensitivity in detecting DR by teleophthalmology compared to the gold standard of an in-person dilated fundus examination ^24^. Since the fundus photographs acquired in our teleophthalmology system were limited to showing the macula, optic nerve, and temporal arcades, DR-related pathology in the peripheral retina could have been missed, though vision-threatening retinopathy was likely to have been captured in fundus images. Previous studies have suggested that single-field fundus photography is sensitive and specific for diagnosing diabetic retinopathy^15^. Additionally, among patients in our study who followed up at BEIS, the concordance between in-person examination and teleophthalmology for detecting the presence or absence of DR was 82.4%. Among patients with ungradable images, a similar rate of DR and DME were identified in eyes that had an in-person examination at BEIS compared to the rate diagnosed in our teleophthalmology-graded patients. Furthermore, a subset of patients who presented for consecutive annual teleophthalmology examinations showed concordance of disease between the two examinations.

The teleophthalmology system served a wide demographic range of patients in terms of age, gender, race, and ethnicity reflecting the diverse communities in which the clinics were located. Interestingly, self-reported non-White patients were more than twice as likely to have an ungradable image, a trend that was reflected across all non-White ethnic groups and does not appear to be explained by differences in age since the median age of non-White patients was younger than White patients, and ungradable patients across all racial and ethnic groups tended to be older. Ungradable fundus images were almost always too dark, which could arise from poor alignment of the eye’s visual axis with the camera’s imaging axis, small pupil, cataract or other media opacity, or increased pigmentation of the fundus. BEIS specialists assessing images were not aware of the patient’s self-reported race or ethnicity. Additional investigation is needed in this area to enhance equitable performance of the teleophthalmology system across demographic groups.

Primary care practices widely employ the HEDIS performance measures as both an internal assessment of quality of care and to qualify for reimbursement and operational incentives. Prior to implementation of the teleophthalmology system, the primary care sites for which HEDIS data was accessible had a 49.5% average of diabetic patients who had an annual eye exam. This measure increased across all sites for the duration of the teleophthalmology program, which was initiated at four out of five primary care sites two months before the state of California enacted a shelter-at-home policy in response to COVID-19. In spite of challenges to healthcare access during the COVID-19 pandemic, the percentage of diabetic patients receiving an eye exam increased 17.1% above baseline. By the end of the phase of the teleophthalmology program reported here, that percentage reached 73.2%, well above the 90th percentile HEDIS goal of 67.89% for which the organization had aimed. During the same interval, two other Stanford-affiliated primary care sites in the Bay Area that did not implement the teleophthalmology system decreased in the same HEDIS measure by 2.3%. In addition to the percentage of patients screened, both the upward trend among sites that implemented teleophthalmology and the downward trend among sites that did not implement teleophthalmology were also reflected in the absolute number of diabetic patients receiving an annual eye examination.

Although the teleophthalmology system likely contributed to this improvement, two aspects of the data suggest that there were other factors contributing to this trend. First, prior to implementation of the tele-ophthalmology system, all of the participating sites demonstrated a gradual upward trend in the HEDIS measure. Second, the number of patient encounters in the teleophthalmology program represents only 3.85% of all diabetic patients comprising the HEDIS measure over the same time period. The additional 13% improvement may have come from other initiatives within the practices to improve HEDIS measures or other factors outside of the healthcare system not accounted for in this study. For instance, the participating primary care sites had concurrent outreach initiatives for traditional ophthalmic screening to fulfill value-based care arrangements. Nevertheless, the number of patients that the teleophthalmology system served could have potentially mitigated the loss in diabetic eye exams by the non-participating sites, both of which were already operating at high HEDIS scores at baseline.

Furthermore, the teleophthalmology system reported here represents a pilot phase of the program. With additional scaling across primary care sites and additional coverage of patients within each practice, the system could achieve greater impact. We are additionally exploring the use of artificial intelligence algorithms to provide even greater access to specialist-level diabetic eye screening.

## CONCLUSIONS

In summary, we saw that primary care-based teleophthalmology improves access to eye exam for diabetic patients and identifies patients with diabetic retinopathy across diverse communities. The use of teleophthalmology has the potential to have far-reaching impacts through greater patient coverage and should be further investigated for larger-scale implementation of eye care services.

## Data Availability

All data produced in the present study are available upon reasonable request to the authors.

## LIST OF ABBREVIATIONS

DR: diabetic retinopathy
DME: diabetic macular edema
BEIS: Byers Eye Institute at Stanford
HEDIS: Healthcare Effectiveness Data and Information Set

## ETHICS APPROVAL AND CONSENT TO PARTICIPATE

The study was approved by the institutional ethics board of Stanford University and conducted in accordance with the tenets of the Declaration of Helsinki. Individual consent for this retrospective analysis was waived. Not applicable.

## CONSENT FOR PUBLICATION

Not applicable.

## AVAILABILITY OF DATA AND MATERIALS

All data generated or analyzed during this study are included in this published article and its supplementary information files.

## COMPETING INTERESTS

The authors state the following competing interests:

E.R.D - no competing interests

K.C. - no competing interests

N.C.K. - no competing interests

C.P. - no competing interests

D.M. - no competing interests

K.M. – no competing interests

T.L. - no competing interests

P.M. - no competing interests

D.D. - no competing interests

V.B.M. - no competing interests

J.D. - no competing interests

K.W. - no competing interests

M.K. - no competing interests

A.P. - no competing interests

M.L. - no competing interests

M.B. - no competing interests

M.T. - no competing interests

## FUNDING

The study was funded by an unrestricted grant from Research to Prevent Blindness and a P30 NIH grant to the Byers Eye Institute of Stanford University.

## AUTHOR’S CONTRIBUTIONS

E.D. and K.C. contributed equally to this study. E.D., K.C., N.K., C.P., and D.M. conceptualized and wrote the main manuscript text. K.C. conducted the data analysis and prepared the figures. T.L., P.M., D.D., V.B.M., J.D., K.W., M.K., A.P., M.L., M.B., and M.T. performed organization and administration of the program, and patient care activities and documentation related to the study. All authors read and approved the final manuscript. All authors reviewed the manuscript.

## ACKNOWLEDGEMENTS

The authors wish to acknowledge the nurses, technicians, and managers at our participating primary care sites who facilitated the teleophthalmology program.

---

**Supplemental Figure 1.**
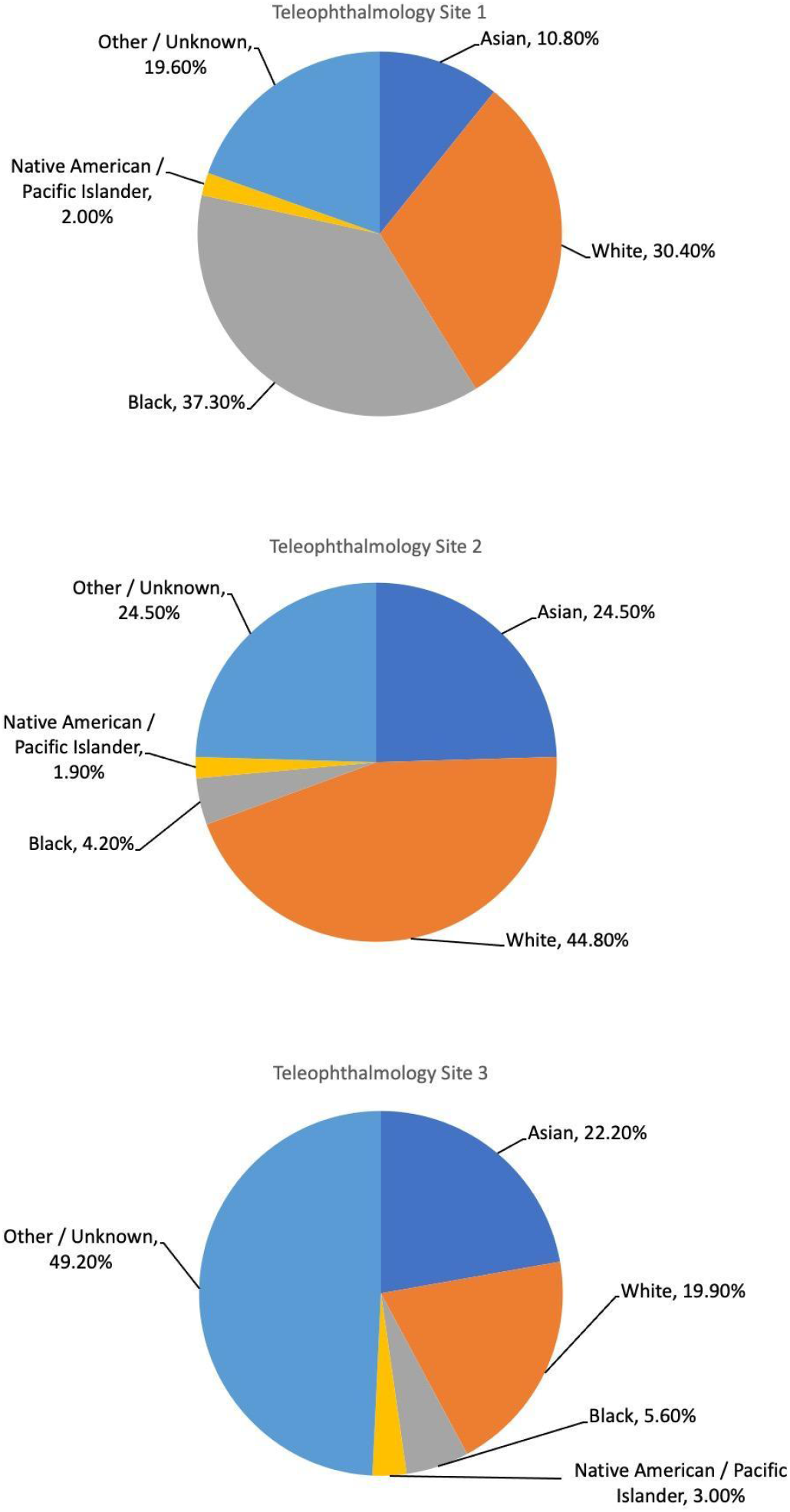
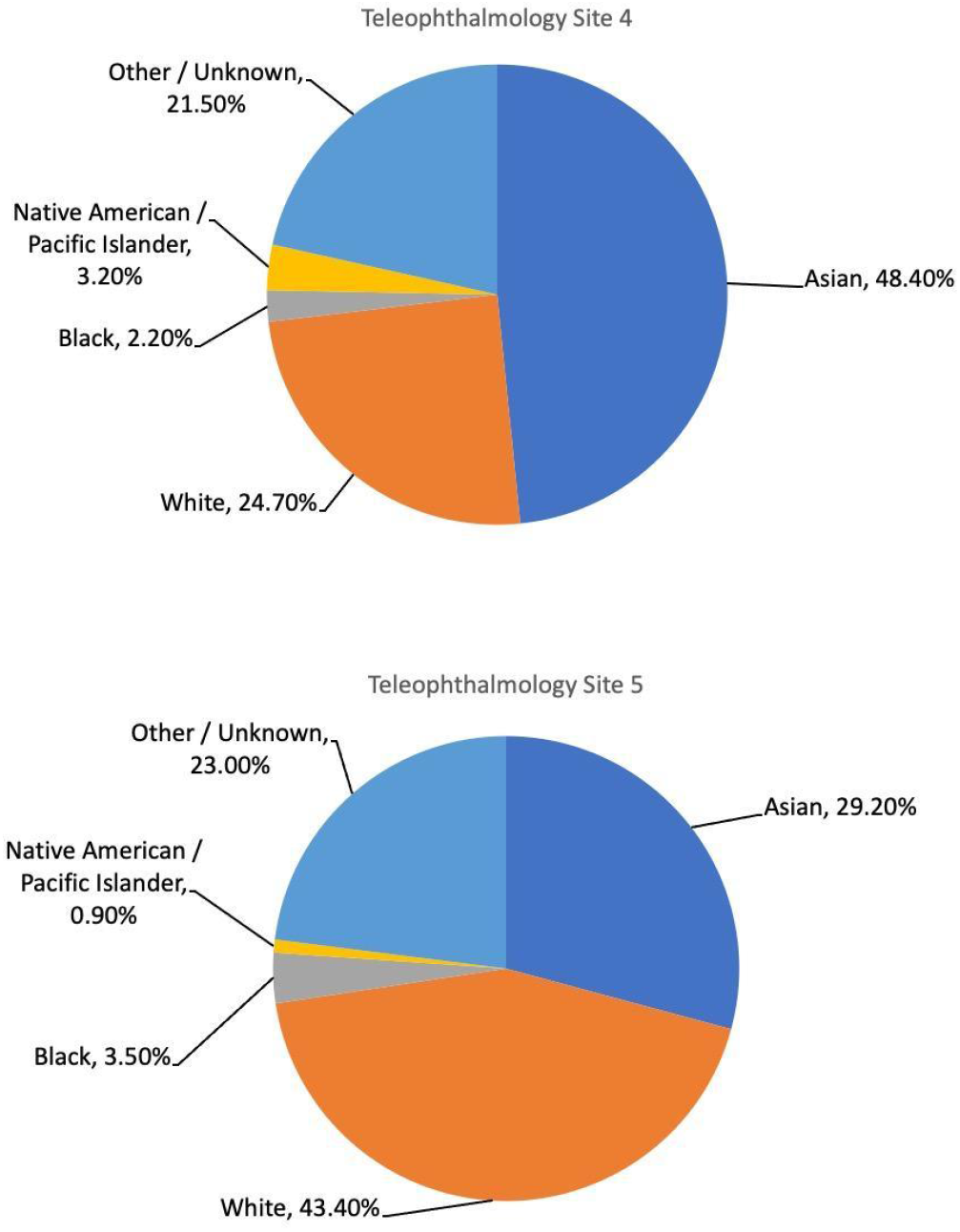
Self-reported race of patients participating in teleophthalmology program per primary care site

## Notes

### Competing Interest Statement

The authors have declared no competing interest.

### Author Declarations

Ethics committee/IRB of Stanford University School of Medicine gave ethical approval for this work.

